# Advancements in Multilingual Biomedical Natural Language Processing: Exploring Large Language Models for Named Entity Recognition and Linking

**DOI:** 10.64898/2026.01.22.26344605

**Authors:** Sara Mazzucato, Tom M. Seinen, Sara Moccia, Silvestro Micera, Andrea Bandini, Erik M. van Mulligen

**Affiliations:** Biorobotics Institute, Department of Excellence in Robotics and AI, Scuola Superiore Sant’Anna, Pisa 56127, Italy; Department of Medical Informatics, Erasmus University Medical Center, Rotterdam 3015 CN, The Netherlands; Health Science Interdisciplinary Research Center, Scuola Superiore Sant’Anna, Pisa 56127, Italy; Department of Innovative Technologies in Medicine and Dentistry, Università degli Studi “G. d’Annunzio”, Chieti, Pescara 66100, Italy; Department of Engineering “Enzo Ferrari”, University of Modena and Reggio Emilia, Modena, Italy; Bertarelli Foundation Chair in Translational Neural Engineering, Center for Neuroprosthetics and Institute of Bioengineering, Ecole Polytechnique Fédérale de Lausanne, Lausanne 1015, Switzerland

**Keywords:** Clinical natural language processing, Named entity recognition, Entity linking, Unified Medical Language System, Multilingualism, Large language models, Electronic health records, Disorders

## Abstract

**Background:** Clinical natural language processing tools are concentrated on English, and rigorous multilingual evaluations are scarce. We address the question of how closely a single zero-shot, prompt-based large language model (LLM) pipeline, with no task-specific training, can reproduce a human disorder annotator for named entity recognition and Unified Medical Language System (UMLS) concept linking across English, Dutch, and Italian.

**Methods:** An English disorder corpus (ShARe/CLEF eHealth dataset, with gold clinician annotations) was ported to Dutch and Italian by an annotation-preserving translation that embeds entity markers before translation, giving aligned multilingual benchmarks. GPT-4o extracted disorder mentions zero-shot; extracted spans were linked to UMLS concepts by embedding similarity. Recognition was scored under both overlap and exact-boundary (strict) criteria, and linking under both a corpus-restricted and a realistic, semantic-type-filtered UMLS search space, with 95% bootstrap confidence intervals, significance testing, and dictionary, learning-based, and zero-shot baselines.

**Results:** Under the overlap criterion, recognition reached F1 0.60 (English), 0.63 (Dutch), and 0.65 (Italian), with high precision (0.73–0.83) and lower recall; on clinically linkable disorders F1 was 0.63 to 0.70. In English the LLM was competitive with established clinical taggers (MedCAT, QuickUMLS; F1 0.63–0.64) at higher precision, and well above a zero-shot GLiNER baseline (0.11–0.19). Strict-boundary recognition lowered F1 to 0.48–0.52, and moving from corpus candidates to a realistic UMLS linking space lowered accuracy@1 from 0.90–0.94 to 0.56–0.69 (best with multilingual SapBERT); both show that reported performance depends strongly on evaluation design. Most false positives were valid clinical concepts (boundary variants or unannotated findings) rather than hallucinations.

**Conclusions:** A single zero-shot LLM pipeline reproduces a human disorder annotator moderately and consistently across three languages without annotated training data, remaining below task-trained systems. The main contribution is a reproducible multilingual evaluation that quantifies how span-matching and candidate-space choices shape conclusions. Because the non-English results are obtained on translated benchmark text, validation on native clinical notes and additional entity types is required; all code and predictions are released.

## 1 Background

In recent years, Electronic Health Records (EHRs) have become a cornerstone of real-world data in observational research, providing critical insights into disease patterns, treatment responses, and patient trajectories [1, 2]. While structured data elements such as diagnostic codes, laboratory results, and medication prescriptions are frequently analyzed, a substantial portion of clinical knowledge remains embedded in unstructured free-text entries like clinical notes [3, 4]. These narrative elements often contain nuanced and context-rich information that structured fields cannot fully capture, making them a vital target for Natural Language Processing (NLP) techniques [5].

Among the key NLP tasks for clinical text, Named Entity Recognition (NER) and Biomedical Entity Linking (BEL) are fundamental for transforming raw narratives into structured knowledge representations [6]. However, concept extraction tools in non-English languages remain sparse and often lack rigorous evaluation, especially in languages with limited annotated resources such as Dutch and Italian [7, 8]. This scarcity is particularly problematic in the context of international initiatives that aim to standardize and use health data across countries, such as the European Medical Information Framework (EMIF) [9], European Health Data and Evidence Network (EHDEN) [10], and Data Analysis and Real World Interrogation Network (DARWIN EU) [11]. These projects rely on frameworks like the Observational Medical Outcomes Partnership Common Data Model (OMOP-CDM) [1] to integrate heterogeneous data, and extending NLP capabilities to multiple languages is crucial for their success.

One of the main barriers to expanding these tools is the lack of annotated corpora in non-English languages. Creating such datasets is resource-intensive, requiring expert annotators and domain-specific expertise [12]. Additionally, machine translation alone is insufficient for porting annotated corpora, since word and phrase reordering often misaligns entity positions, leading to annotation loss. Recent work has shown that embedding entity annotations directly in the text before translation, then extracting them post-translation, can mitigate these issues [13].

In this context, our study investigates how prompt-based learning with LLMs can support NER and BEL tasks in Dutch and Italian biomedical texts. We translated a well-established English corpus, the ShARe corpus (SemEval-2014 Task 7) [14], into Dutch and Italian using an annotation-preserving protocol, and evaluated multilingual LLMs under a zero-shot configuration. By integrating BEL capabilities after this prompt-based framework, our pipeline offers a lightweight yet flexible alternative to traditional toolchains, contributing to resource-efficient multilingual NLP for clinical information extraction.

**Contributions.** This work provides an empirical, reproducible evaluation of zero-shot multilingual clinical concept extraction. Specifically, we offer: (i) the first head-to-head assessment, to our knowledge, of a zero-shot LLM recognition plus embedding-linking pipeline for *disorder* extraction across English, Dutch, and Italian on a shared benchmark; (ii) a controlled analysis of how evaluation choices (overlap vs. exact-boundary matching, restricted vs. realistic UMLS candidate space) change apparent performance; (iii) a comparison against clinical and open-weight baselines (MedCAT, QuickUMLS, GLiNER, SapBERT) and the published SemEval-2014 Task 7 results [14]; and (iv) a quantitative, per-category error analysis of where the pipeline helps and where it fails. The components (annotation-preserving translation, zero-shot extraction, embedding-based linking) build on prior work, so the contribution is this comparative evaluation.

### 1.1 Related Work

Generative Large Language Models (LLMs) such as GPT-4 have recently emerged as promising tools for multilingual information extraction and corpus generation [15]. Their ability to perform zero-shot prompting opens opportunities for building concept extraction pipelines without requiring extensive annotated data [16, 17]. While LLM-based systems have begun to be explored for clinical NLP in English [18], their performance in multilingual settings remains largely underexplored.

Discriminative transformer-based models such as BERT [19] and its domain-specific adaptations like BioBERT [20] and SapBERT [21] remain highly effective for entity linking tasks. These models generate dense vector representations that allow for efficient semantic matching between mentions and knowledge base concepts. Recent works have shown that contrastively trained biomedical embeddings can outperform generative models on structured entity linking benchmarks [21].

Tools like MetaMap [22], QuickUMLS [23], and MedCAT [24] have shown strong performance in English, particularly when grounded in terminologies like the Unified Medical Language System (UMLS). Extending these methods to languages with limited annotated resources requires innovative translation strategies combined with robust model adaptation [7, 8].

Beyond English, community shared tasks have driven clinical NER and entity linking in other languages, most notably the Spanish series organised within BioASQ/CLEF: DisTEMIST for disease detection and normalisation [25], together with MedProcNER (clinical procedures), SympTEMIST (symptoms and multilingual linking) [26], and CodiEsp (ICD-10 coding). These efforts show that high-quality, natively annotated non-English clinical corpora are achievable and provide the kind of realistic, full-search-space linking evaluation we adopt here. On the modelling side, lightweight zero-shot recognisers such as GLiNER [27] and rule/ML clinical toolkits such as medspaCy [28] offer multilingual or language-portable baselines against which prompt-based LLM extraction should be measured. For linking, contrastively trained encoders such as SapBERT [21] remain the reference approach. We therefore compare our pipeline against SapBERT for linking and GLiNER for zero-shot NER, and additionally against the published SemEval-2014 Task 7 shared-task results [14].

International initiatives such as EMIF, EHDEN, and DARWIN EU further emphasize the need for multilingual NLP capabilities, highlighting the importance of annotation-preserving strategies for corpus translation [9–11]. This body of work motivates the design of our pipeline, which leverages the complementary strengths of generative and discriminative models for NER and BEL in Dutch and Italian clinical texts.

## 2 Methods

### 2.1 Data and Setting

The experimental setup consisted of three main steps: corpus translation and preparation, concept recognition, and entity linking, as illustrated in Figure 1. We used the disorder corpus of SemEval-2014 Task 7 (Analysis of Clinical Text), which was also run as Shared Task 2 of ShARe/CLEF eHealth 2014, hereafter the SC corpus [14]. The corpus comprises 430 de-identified clinical notes (299 for training and 131 for testing; 131 test notes were retained for evaluation, see Corpus Statistics) of several types (discharge summaries and echocardiogram, electrocardiogram, and radiology reports) drawn from the MIMIC II database, with disorder mentions annotated and normalized to UMLS Concept Unique Identifiers (CUIs) in the SNOMED-CT subset. The corpus was transformed into a tabular format, separating text documents (attributes: DocumentId, Text) and concept annotations (attributes: DocumentId, CUI, SpanStart, SpanEnd, SpanText).

**Figure 1:**
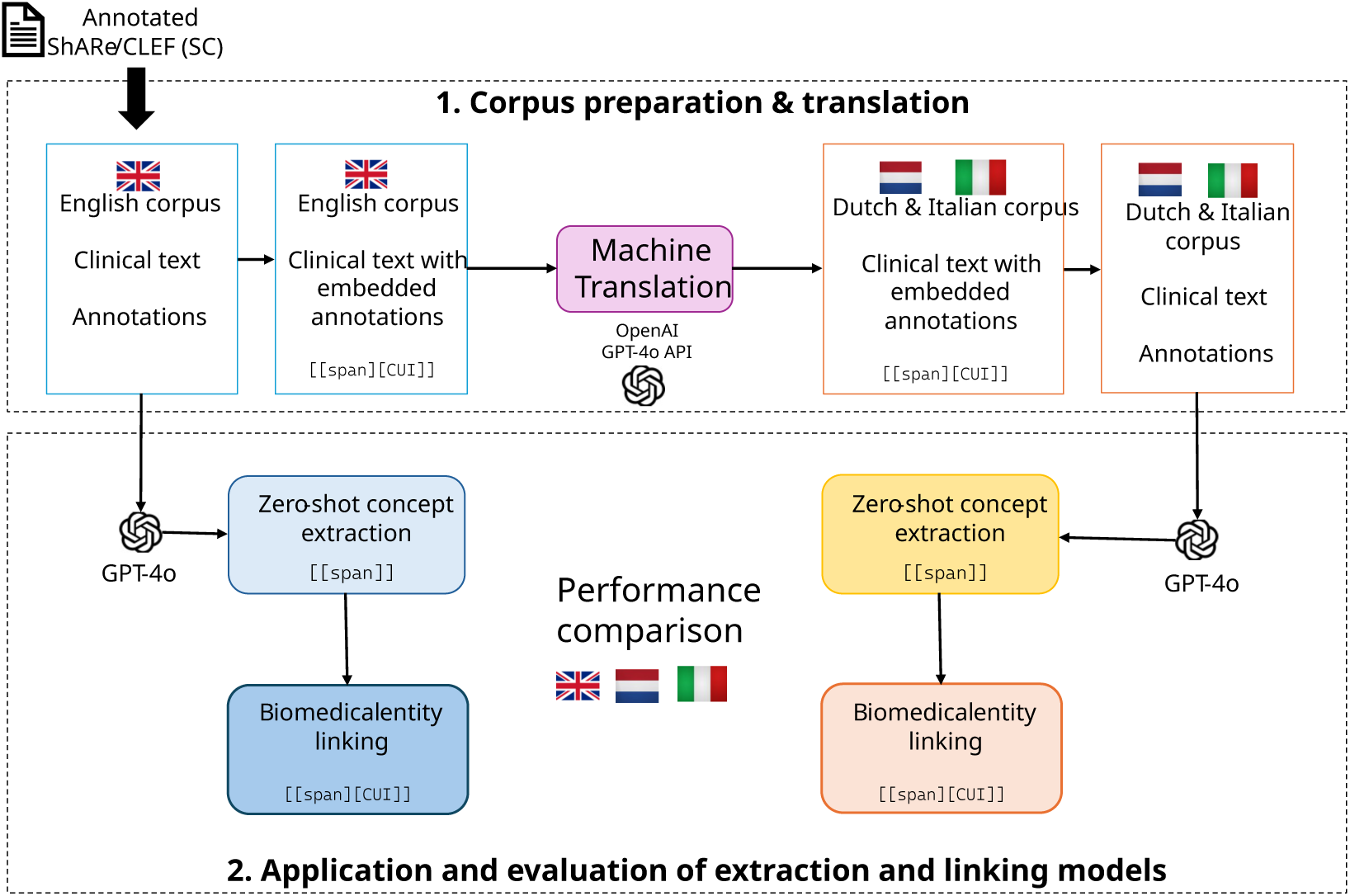
Experimental setup: 1. preparation and translation of the dataset, and 2. appli­cation and evaluation of an LLM for named entity recognition followed by biomedical entity linking of the recognized entities. CUI: concept unique identifier.

#### Normalizable and CUI-linkable mentions

In the gold standard each disorder mention is either *CUI-bearing*, i.e. normalized by the corpus annotators to a UMLS CUI in the SNOMED-CT subset, or *CUI-less*, i.e. marked as a disorder but not mappable to a single UMLS concept and recorded with the placeholder code C0000000. CUI-less mentions include some negated or normal physical-exam findings and abbreviated fragments, and make up about a quarter of the English and Italian gold and none of the Dutch gold. We refer to the CUI-bearing subset as the *CUI-linkable gold*. Recognition is reported both against the *full gold* (all disorder mentions) and against the *CUI-linkable gold* (CUI-bearing mentions only); in the latter, predicted spans that overlap a CUI-less gold mention count as neither correct nor incorrect. Because a linker cannot assign a valid concept to a CUI-less mention, all entity-linking metrics are computed on the CUI-linkable subset. Code to reproduce the experiments is publicly available at https://github.com/saramazz/llm_EMC.

### 2.2 Corpus Translation

The dataset was processed through a multi-step pipeline to standardize both the raw clinical text and its annotations (Figure 2). The corpus translation method involved embedding annotations within the text prior to translation, followed by extraction post-translation, ensuring linguistic and conceptual accuracy were maintained [13].

**Figure 2:**
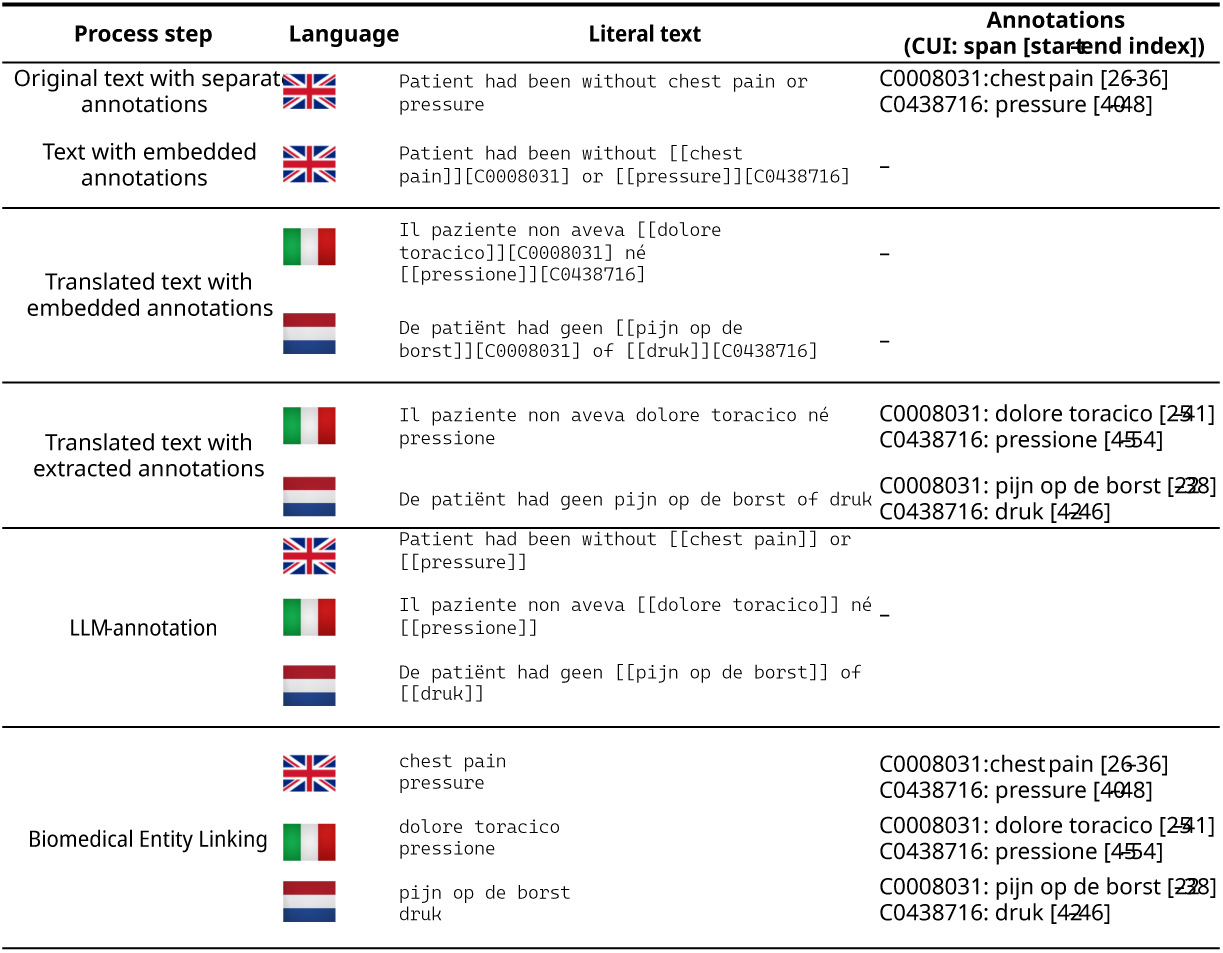
Illustrative example of the workflow applied to a sample clinical note. To enable accurate translation without losing annotation alignment, annotated spans were embedded directly into the clinical text using inline markers of the form [[span][CUI]], thus preserving annotation boundaries within the text. Then, named entity recognition was performed using GPT-4o with a structured prompt tailored to identify biomedical entities while maintaining the integrity of the translated text. Finally, biomedical entity linking allowed to link the recognized entities to CUIs.

Translation was performed using OpenAI’s GPT-4 Turbo API in a zero-shot setting, prompted as follows:

*Translate the document to <target language>. Keep the formatting the same, including the in-text annotations: [[span][code]]*.

Annotations were extracted from the translated text using the regular expression pattern “\[\[([^\]\[]\ast )\]\[(C[0–9]\ast )\]\]” to recover span and CUI pairs despite linguistic shifts.

The annotation-preserving protocol follows Seinen et al. [13]. The BLEU [29] (0.46, 0.58) and chrF [30] (0.75, 0.87) scores reported in that work compare GPT and Google-Translate outputs against each other rather than against a clinician reference translation.

#### 2.2.1 Corpus Statistics

Table 1 summarises the corpus across language splits. The three splits are comparable in length, with a mean note length of 1,190 to 1,317 tokens and Italian notes the longest; only 3 to 6 notes per language exceed 2,000 tokens.

**Table 1:** Corpus-level statistics by language split. Length and vocabulary statistics are computed on the test corpus. “Annotations projected” reports the share of English CUI-bearing gold mentions recovered in the same document after translation and back-projection. Counts are over the 131-document evaluation set. The Dutch gold contains no CUI-less mentions, so its CUI-bearing count equals its gold count.

| **Statistic** | **English** | **Dutch** | **Italian** |
| --- | --- | --- | --- |
| Documents (test split) | 131 | 131 | 131 |
| Tokens | 160,867 | 155,865 | 172,486 |
| Mean note length (tokens) | 1,228 | 1,190 | 1,317 |
| Notes > 2,000 tokens | 3 | 3 | 6 |
| Vocabulary (unique tokens) | 9,619 | 11,911 | 11,028 |
| Mean mention length (chars) | 11.2 | 11.8 | 12.5 |
| Abbreviations (% of tokens) | 5.0 | 4.9 | 4.4 |
| Disorder mentions (gold) | 7,966 | 7,758 | 7,928 |
| CUI-bearing mentions | 5,973 | 7,758 | 5,938 |
| Annotations projected (%) | n/a | 98.1 | 99.2 |

Translation increases lexical diversity: the Dutch and Italian splits have larger vocabularies (11,911 and 11,028 unique tokens) than English (9,619) at comparable total length, reflecting morphological richness and translation variants. Mean mention length is slightly greater in the translations (11.8 and 12.5 characters versus 11.2), while abbreviations form a similar share of tokens across languages (4.4 to 5.0%); the cross-lingual recall differences reported below are therefore not explained by an aggregate difference in abbreviation density.

### 2.3 Named Entity Recognition

Concept recognition was performed using the GPT-4o large language model. GPT-4o was chosen as a strong, widely available general-purpose LLM with documented zero-shot multilingual capability, allowing a single prompt to be applied unchanged across English, Dutch, and Italian. At the time of the experiments it was among the strongest models for multilingual clinical text, and using one hosted model avoided the per-language fine-tuning that supervised taggers require. To support reproducibility, we release our saved predictions and evaluation code and include an open-weight extractor (GLiNER) as a reference point. The pipeline loads unannotated clinical notes from language-specific corpora (English, Dutch, Italian) and processes each note in parallel. Prompt-based requests to GPT-4o used a maximum of 2000 tokens per request. The prompt template was based on the SemEval-2014 Task 7 annotation guidelines; the full prompt is provided in Additional file 1 (Section S1).

Entity markers were removed before extraction, so GPT-4o received unannotated text. Because the same GPT-4 model family produced both the translated corpora and the extractions, we ran an open-weight extractor (GLiNER [27]) on the same text as a leakage control and compared against the published SemEval-2014 Task 7 results [14]; we interpret these controls in the Limitations.

### 2.4 Biomedical Entity Linking

BEL was performed by mapping the recognized entities to UMLS CUIs [31] using embedding similarity. The recognized entities were lowercased before embedding and abbreviations were left unexpanded and no punctuation removal was applied, so that all three languages share an identical setting, since a validated UMLS abbreviation inventory exists for English but not for Dutch or Italian. A post-hoc English analysis shows this is conservative: expanding abbreviations with a training-set most-frequent-sense rule would raise top-1 linking from 0.73 to 0.97.

#### Candidate generation

We report two candidate-space settings. In the *restricted* setting, mentions are matched only against the set of CUIs that occur as gold annotations in the corpus [13]; this is an upper-bound condition that does not reflect deployment. In the *realistic* setting, mentions are matched against a UMLS 2024AB subset restricted to clinically relevant (disorder) semantic types, comprising 148,134 (English), 58,406 (Dutch), and 30,064 (Italian) concepts, represented by 727,198 / 127,045 / 46,770 name–synonym strings respectively. Because some gold CUIs fall outside this subset, accuracy under the realistic setting is bounded above by 0.85 (English), 0.88 (Dutch), and 0.77 (Italian). The disorder subset is built from all UMLS name and synonym strings of the retained concepts, which include the abbreviation and acronym atoms UMLS records for disorders (for example “MI” for myocardial infarction), so abbreviated disorder mentions can still match a concept; abbreviations that denote non-disorders (for example “ECG”, a diagnostic procedure) fall outside the disorder-only gold and are therefore out of scope.

#### Vector database

Candidate strings and their CUIs were stored in a local ChromaDB vector database [32]. Indexes were *language-specific*: mentions in each language were matched only against UMLS atoms in the same language (the realistic indexes held 727,198 English, 127,045 Dutch, and 46,770 Italian entries; the restricted indexes held the corpus-CUI strings, roughly 2,500–3,300 entries per language). During inference each mention was embedded and compared by cosine similarity, retrieving the top-5 CUIs to mitigate embedding/score noise, capture alternative senses of ambiguous mentions, and provide candidates for subsequent context-based disambiguation or filtering.

#### Embedding models

Three encoders were compared: a lightweight general-purpose sentence encoder (all-MiniLM-L6-v2), a biomedical model (BioBERT), and a domain-specific multilingual entity-linking encoder (multilingual SapBERT, SapBERT-UMLS-2020AB-all-lang-from-XLMR) [21]. all-MiniLM was included as a strong lightweight general baseline, BioBERT as a widely used biomedical model, and multilingual SapBERT because it is the reference contrastively trained encoder for biomedical entity linking and natively covers English, Dutch, and Italian. Mentions and candidates were embedded with the same model and compared by cosine similarity (CLS pooling with L2 normalisation for SapBERT).

### 2.5 Evaluation

NER performance was assessed with precision, recall, and F1 under two criteria. Following Seinen et al. [13], the *overlap* criterion (any character overlap between a predicted and a gold span within the same document) is the primary metric and measures mention-level agreement with the human annotator; the *strict* criterion (exact boundary match) is reported alongside for comparability with the clinical NLP literature [33]. Matching is robust to line-number shifts between the GPT output and the gold (content-aligned lines with a token-overlap guard).

BEL performance was evaluated using accuracy@*{*1,3,5*}* and mean reciprocal rank (MRR) [34], reported for both the restricted and the realistic candidate spaces defined above; because each mention has a single gold CUI, precision@1, recall@1 and F1@1 coincide with accuracy@1. Recognition is reported against both the full gold and the CUI-linkable gold defined in the Data and Setting section.

#### Uncertainty

We report 95% confidence intervals for all primary metrics from 1,000 bootstrap resamples over documents, and test between-language differences with Mann–Whitney U tests under Holm correction for multiple comparisons.

## 3 Results

Across English, Dutch and Italian, the zero-shot pipeline reproduced the human disorder annotator moderately, with recognition overlap F1 of 0.60, 0.63 and 0.65, respectively, at high precision and lower recall (Tables 2 and 3). How favourable these numbers appear depends on two evaluation choices: for recognition, overlap versus strict (exact-boundary) matching and the full versus the CUI-linkable gold; for linking, a restricted corpus-CUI candidate set versus a realistic, full-UMLS search space, alongside dictionary, learning-based, and zero-shot baselines. Span *overlap* is the primary matching criterion (mention-level agreement with the human annotator), and recognition on the CUI-linkable gold is computed over CUI-bearing mentions only, as defined in the Data and Setting section.

**Table 2:**
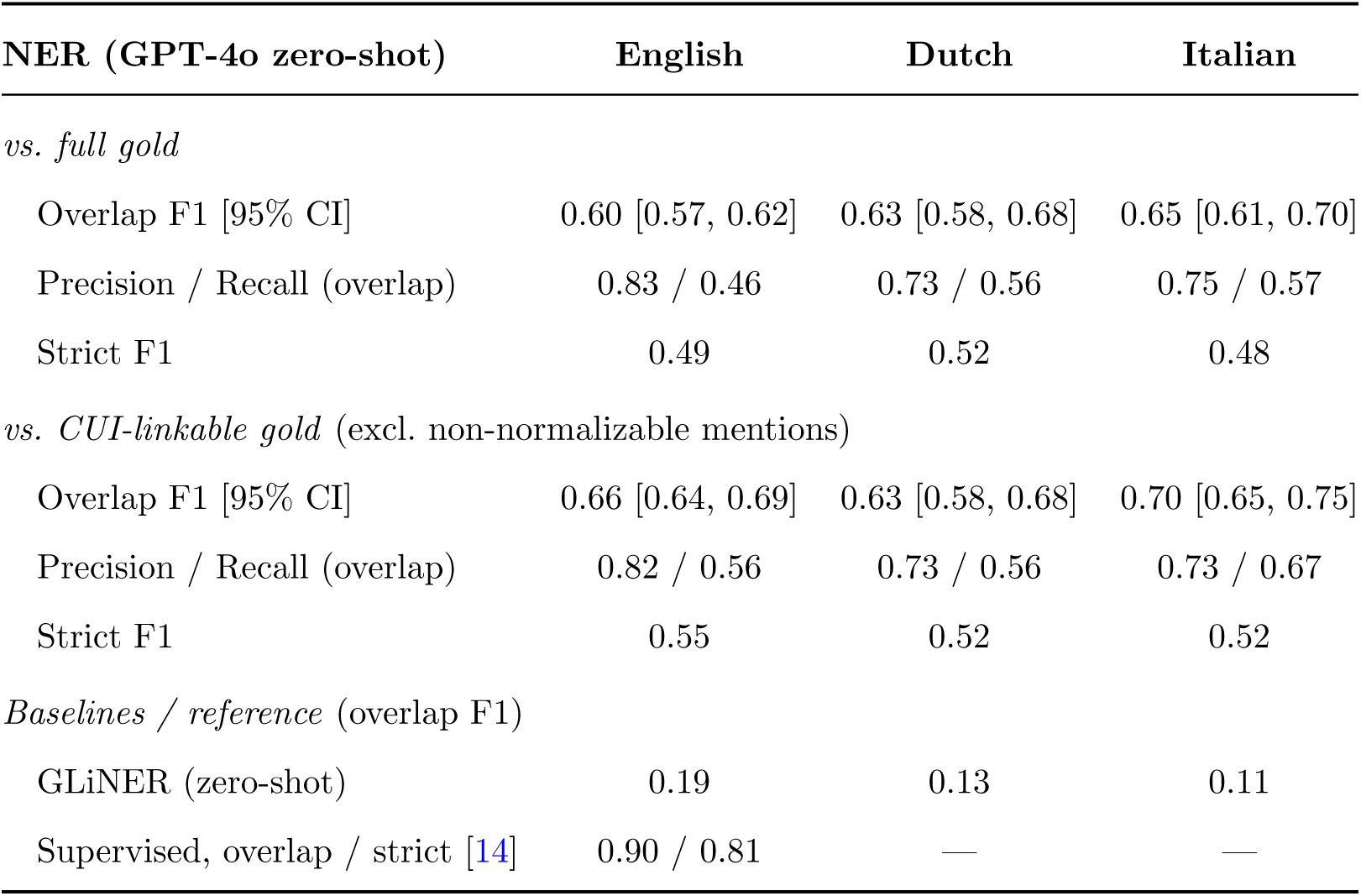
GPT-4o zero-shot NER by language, as agreement with the gold annotator under overlap (primary) and strict (exact-boundary) matching, with 95% bootstrap confidence intervals, for the full gold and the CUI-linkable gold (CUI-less mentions, about a quarter of the English and Italian gold and none for Dutch, count as neither correct nor incorrect). Baselines: zero-shot GLiNER and the best supervised SemEval-2014 Task 7 system (UTH CCB).

| NER (GPT-4o zero-shot) | English | Dutch | Italian |
| --- | --- | --- | --- |
| *vs. full gold* |  |  |  |
| Overlap F1 [95% CI] | 0.60 [0.57, 0.62] | 0.63 [0.58, 0.68] | 0.65 [0.61, 0.70] |
| Precision / Recall (overlap) | 0.83 / 0.46 | 0.73 / 0.56 | 0.75 / 0.57 |
| Strict F1 | 0.49 | 0.52 | 0.48 |
| *vs. CUI-linkable gold* (excl. non-normalizable mentions) |  |  |  |
| Overlap F1 [95% CI] | 0.66 [0.64, 0.69] | 0.63 [0.58, 0.68] | 0.70 [0.65, 0.75] |
| Precision / Recall (overlap) | 0.82 / 0.56 | 0.73 / 0.56 | 0.73 / 0.67 |
| Strict F1 | 0.55 | 0.52 | 0.52 |
| *Baselines / reference* (overlap F1) |  |  |  |
| GLiNER (zero-shot) | 0.19 | 0.13 | 0.11 |
| Supervised, overlap / strict [14] | 0.90 / 0.81 | — | — |

**Table 3:** Biomedical entity linking by language: accuracy@1 for the restricted and realistic UMLS candidate sets (realistic value in parentheses on linkable mentions), and mean reciprocal rank (restricted / realistic). The candidate-coverage ceiling is the maximum reachable accuracy@1. mSapBERT, multilingual SapBERT.

| **BEL accuracy@1** | **English** | **Dutch** | **Italian** |
| --- | --- | --- | --- |
| *Restricted candidate set* |  |  |  |
| all-MiniLM-L6-v2 | 0.94 | 0.91 | 0.90 |
| BioBERT | 0.94 | 0.91 | 0.90 |
| mSapBERT (multilingual) | 0.94 | 0.91 | 0.91 |
| *Realistic UMLS set* (linkable subset in parentheses) |  |  |  |
| all-MiniLM-L6-v2 | 0.69 (0.77) | 0.56 (0.56) | 0.52 (0.60) |
| BioBERT | 0.26 (0.29) | 0.49 (0.49) | 0.35 (0.40) |
| mSapBERT (multilingual) | 0.69 (0.77) | 0.63 (0.63) | 0.56 (0.64) |
| Candidate-coverage ceiling | 0.85 | 0.88 | 0.77 |
| *Mean reciprocal rank* (restricted / realistic) |  |  |  |
| all-MiniLM-L6-v2 | 0.96 / 0.72 | 0.93 / 0.59 | 0.93 / 0.55 |
| BioBERT | 0.95 / 0.32 | 0.91 / 0.52 | 0.92 / 0.38 |
| mSapBERT (multilingual) | 0.95 / 0.72 | 0.93 / 0.65 | 0.93 / 0.59 |

The NER agreement with the gold annotator is visible in Figure 3a. Against the full gold, overlap F1 was 0.60 in English (95% CI 0.57–0.62), 0.63 in Dutch (0.58–0.68), and 0.65 in Italian (0.61–0.70); precision was high and highest in English (0.83, vs 0.73 Dutch and 0.75 Italian) while recall was lower (0.46–0.57), indicating a conservative model that tags fewer mentions but is usually correct. Under the strict exact-boundary criterion F1 was 0.49 (English), 0.52 (Dutch), and 0.48 (Italian). Restricting to CUI-linkable gold (excluding the non-normalizable mentions, about a quarter of the English and Italian gold; Dutch has no CUI-less mentions) raised overlap F1 to 0.66 in English and 0.70 in Italian; Dutch is unchanged at 0.63. For external context, the best *supervised* system in the SemEval-2014 Task 7 disorder-recognition task reached a relaxed (overlap) F1 of 0.90 and a strict F1 of 0.81 [14], both well above the zero-shot pipeline (overlap 0.60, strict 0.49), confirming that zero-shot extraction remains below trained systems; a zero-shot GLiNER baseline scored far lower still (overlap F1 0.19, 0.13, 0.11).

**Figure 3:**
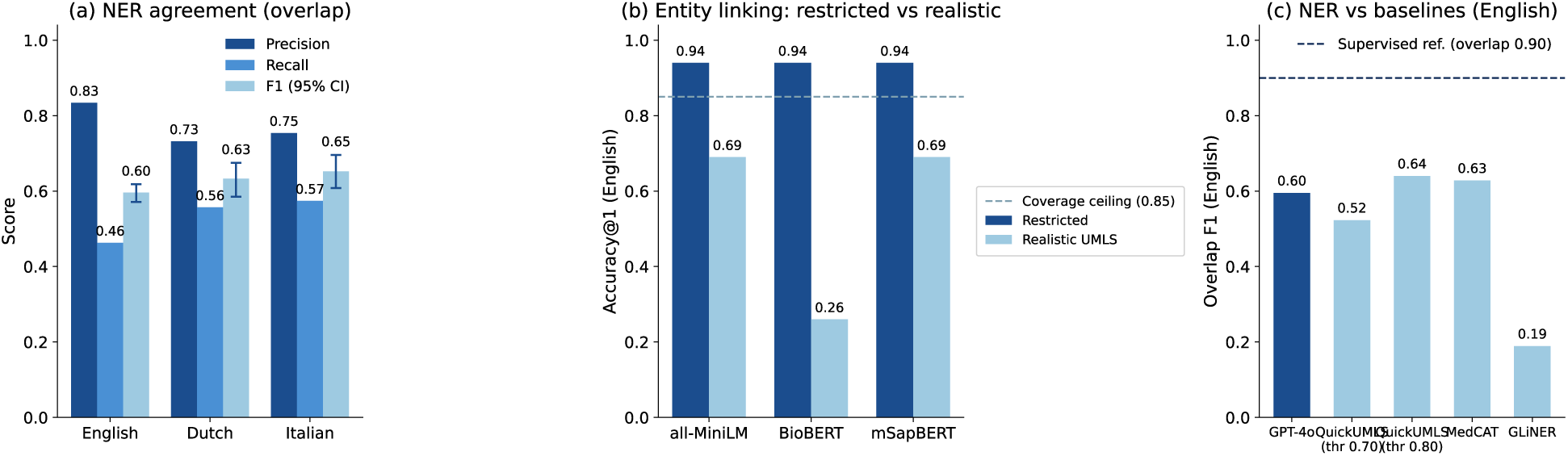
Performance across English, Dutch and Italian on the ShARe/CLEF (SC) cor­pus. (a) GPT-4o NER agreement with the gold annotator under overlap matching, showing precision, recall and F1 by language; error bars are 95% bootstrap confidence intervals on F1. (b) Biomedical entity linking accuracy@1 by encoder in the restricted versus realistic, semantic-type-filtered UMLS candidate setting (English); the dashed line is the candidate-coverage ceiling. (c) English NER: GPT-4o compared with clinical concept-extraction tools (MedCAT, QuickUMLS at two similarity thresholds) and a zero-shot GLiNER baseline, over­lap F1; the dashed line is the supervised SemEval-2014 Task 7 reference (overlap F1 0.90). Figure generated by `figures_v2.py`.

Against established clinical concept-extraction tools run on the English notes (Table 4), the picture is nuanced. GPT-4o is by far the most precise system (precision 0.83) but also the most conservative, so dictionary- and learning-based taggers that annotate more aggressively reach higher overlap F1 (QuickUMLS up to 0.64 at a stricter threshold, MedCAT 0.63) and higher strict F1 (0.51 and 0.54 versus 0.49), at the cost of much lower precision. MedCAT, documented as trained on MIMIC-III (the source of our corpus), also gives the best concept accuracy (CUI@1 0.75). The zero-shot LLM is therefore competitive with specialised English tools; its distinguishing feature is that the same pipeline transfers unchanged to Dutch and Italian, for which such tools are not available.

**Table 4:** English NER: the zero-shot LLM versus established clinical concept-extraction tools on the same document gold (7,966 disorder mentions). P, R and F1 are overlap-based, strict F1 requires exact boundaries, and CUI@1 is the share of correctly bounded predictions whose CUI matches the gold (over predictions with a normalizable gold CUI). QuickUMLS is shown at thresholds 0.70 (default) and 0.80; its F1 reaches 0.68 at 0.95. MedCAT (`umls_sm`) [24] is documented as trained on MIMIC-III, the source of our corpus, which may favour it. All systems share the same gold and overlap criterion; under the tools’ raw-offset scoring GPT-4o reaches overlap F1 0.56, so the gap is robust to the scorer. These tools are English-only.

| **System** | **n<sub>pred</sub>** | **P** | **R** | **F1** | **Strict F1** | **CUI@1** |
| --- | --- | --- | --- | --- | --- | --- |
| GPT-4o (zero-shot) | 4,421 | 0.83 | 0.46 | 0.60 | 0.49 | n/a |
| QuickUMLS (thr 0.70, default) | 12,595 | 0.43 | 0.68 | 0.52 | 0.34 | 0.66 |
| QuickUMLS (thr 0.80) | 8,156 | 0.63 | 0.65 | 0.64 | 0.51 | 0.74 |
| MedCAT (`umls_sm`) | 10,742 | 0.55 | 0.74 | 0.63 | 0.54 | 0.75 |

A systematic analysis of the matched set clarifies the cross-lingual precision pattern. English produces far fewer false positives (735) than Dutch (1,586) or Italian (1,486), and they are of a different kind: 78% of the English false positives are isolated spans with no overlapping gold mention, typically non-disorder concepts such as procedures (e.g. “hysterectomy”, “transurethral resection”) that the disorder-only gold does not annotate, whereas in Dutch and Italian roughly half (46% and 54%) are boundary variants of a correctly identified concept, the right disorder with a different span extent. The high English precision (0.83) thus reflects both fewer false positives and the fact that they are genuinely foreign to the gold, while the lower Dutch and Italian precision is driven by span-boundary disagreements rather than wrong concepts (a per-semantic-type recall breakdown is given in Table 5).

**Table 5:** GPT-4o recall by UMLS disorder semantic type, on the CUI-bearing gold under over­lap matching (per-type group sizes 143 to 2,541 mentions per language). Disease/Syndrome is the largest group. English recall is lower than Dutch and Italian across all types, and signs/symptoms, pathologic functions and findings are recovered least reliably. The per-type rows cover mentions whose gold CUI maps to one of the listed semantic types (for Dutch, 5,857 of the 7,758 CUI-bearing mentions); the “All CUI-bearing” recall is computed over all CUI-bearing gold and therefore differs from a simple average of the rows above.

| **Semantic type** | **TUI** | **English** | **Dutch** | **Italian** |
| --- | --- | --- | --- | --- |
| Disease or Syndrome | T047 | 0.73 | 0.77 | 0.78 |
| Sign or Symptom | T184 | 0.43 | 0.59 | 0.60 |
| Pathologic Function | T046 | 0.37 | 0.51 | 0.57 |
| Neoplastic Process | T191 | 0.72 | 0.83 | 0.76 |
| Mental/Behavioral Dysfunction | T048 | 0.65 | 0.78 | 0.74 |
| Injury or Poisoning | T037 | 0.39 | 0.57 | 0.53 |
| Finding | T033 | 0.28 | 0.43 | 0.51 |
| Other | — | 0.44 | 0.51 | 0.54 |
| All CUI-bearing | — | 0.56 | 0.56 | 0.67 |

On the false-negative side, 1,993 of the 7,966 English gold mentions (25%) are non-normalizable (CUI-less); these account for 1,630 of the 4,280 English false negatives (38%), chiefly negated or normal physical-exam findings and fragments, with the remainder split between genuine disorder misses and non-disorder terms. At the concept level, part of the span-level misses are recovered elsewhere in the same document, where the model tagged the same gold concept at another mention; this concept-level recovery rises across languages, from 8.9% of the CUI-bearing English misses to 20.4% in Dutch and 29.9% in Italian, typically abbreviation or synonym variants (e.g. gold “HTN” recovered as “hypertension”).

Recall also varies markedly by disorder semantic type (Table 5). Well-defined diseases and neoplasms are recovered best in all three languages (Disease/Syndrome 0.73 to 0.78, Neoplastic Process 0.72 to 0.83), whereas signs and symptoms, pathologic functions and general findings are recovered far less reliably, and most so in English (Sign/Symptom 0.43 in English vs 0.59 in Dutch and 0.60 in Italian; Finding 0.28 vs 0.43 to 0.51). English recall is lower than Dutch and Italian for every type, consistent with the terse-text effect described above. Precision and F1 are not reported per type because false positives cannot be assigned a gold semantic type.

For entity linking, all three encoders performed similarly on the restricted candidate set, where each disorder is matched only against the small set of corpus CUIs: accuracy@1 was 0.90–0.94 across languages (Figure 3b). This setting is an upper bound that does not reflect deployment. Against a realistic, semantic-type-filtered UMLS candidate set, accuracy@1 dropped substantially: for the multilingual SapBERT model to 0.69 (English), 0.63 (Dutch), and 0.56 (Italian), with all-MiniLM-L6-v2 close behind in English (0.69) but lower in Dutch and Italian, and raw BioBERT embeddings far weaker (0.26–0.49). On the linkable subset (excluding CUI-less gold) SapBERT reached 0.77 (English), 0.63 (Dutch), and 0.64 (Italian). Part of the realistic gap is structural: because some gold CUIs are absent from the candidate set, accuracy@1 is capped at 0.85 (English), 0.88 (Dutch), and 0.77 (Italian). The domain-specific multilingual encoder was thus the most reliable, while the lightweight general-purpose encoder remained competitive, especially in English.

Extending the match to the top three and five candidates raised realistic-setting accuracy only modestly (multilingual SapBERT: English 0.69/0.76/0.77, Dutch 0.63/0.67/0.68, Italian 0.56/0.61/0.62 at ranks 1/3/5; full accuracy@*{*1,3,5*}* and MRR in Additional file 1, Table S4). In English most of the reachable gold concepts are already retrieved within the top five (against the 0.85 coverage ceiling), so re-ranking could recover part of the errors, whereas in Dutch and Italian a larger share is not retrieved at all.

## 4 Discussion

The analysis of GPT annotations on clinical notes suggests that GPT struggles with subtle or peripheral descriptors that do not occupy the syntactic focus of a sentence, consistent with prior observations of LLMs in biomedical NLP where contextually weak cues are often overlooked [33, 35].

Many predictions that count as false positives against the gold standard are in fact valid clinical content rather than hallucinations: in English they are predominantly non-disorder concepts such as procedures, which the disorder-only gold does not annotate, and in Dutch and Italian they are predominantly boundary variants of a correctly identified disorder. Predictions matching no UMLS concept are therefore a minority and represent only an upper bound on hallucination. Negation and assertion handling remains relevant but appears to contribute relatively few errors in this setting [36, 37].

These complementary detections suggest the pipeline can support, rather than replace, human curation by surfacing valid concepts that annotators miss, while still requiring filtering and oversight before any use [35, 38].

English had the highest precision (0.83) but the lowest recall, giving a slightly lower full-gold F1 (0.60) than Dutch (0.63) or Italian (0.65). On CUI-linkable disorders English rose to 0.66 and Italian to 0.70, while Dutch stayed at 0.63 (no CUI-less gold). The lower English recall is a text-style effect: the original notes use terse shorthand (e.g. “Trace AR”, “SPIDER ANGIOMATA”) that GPT tends not to tag, whereas the Dutch and Italian translations render these as explicit mentions. This raises recall but also produces more span-boundary disagreements, which explain much of the lower non-English precision. Because the gold-to-gold projection is near-complete (0.98–0.99) and the per-document CUI overlap is high (mean Jaccard 0.98–0.99), this reflects recognition on original versus translated text, not annotation loss; the Dutch and Italian results should therefore be read as evidence on translated benchmark text, not native clinical notes [7, 39].

For entity linking, the candidate space dominated performance. On the restricted corpus-CUI set all encoders exceeded 0.90, but on a realistic UMLS candidate set the multilingual SapBERT model was the most reliable, the lightweight general-purpose encoder was competitive in English yet weaker in Dutch and Italian, and raw BioBERT embeddings degraded sharply [40]. Embedding-based linking is thus viable but sensitive to both the candidate space and the encoder.

Cross-lingual linking gaps are consistent with sparser non-English coverage in biomedical terminologies and with the candidate-coverage ceiling on the realistic set, so a domain-adapted multilingual encoder is preferable for non-English text [39]. Morphology, synonym variability, and translation style likely contribute as well.

Error analysis shows that the nature of the linking errors depends on the candidate space. In the restricted setting the correct CUI is almost always retrieved and usually ranked first (MRR 0.91 to 0.96), so the few mistakes are re-ranking rather than retrieval failures. In the realistic setting, by contrast, extending to the top five recovers only a small additional fraction (Additional file 1, Table S4), so most misses are cases where the gold concept is not retrieved at all, consistent with sparser non-English coverage and the candidate-coverage ceiling. Ontology-aware re-ranking on top of embedding-based retrieval is therefore promising mainly once candidate coverage is adequate [21].

Considering NER and BEL together, the pipeline only partially replicates the human annotator: recognition is moderate and well below supervised systems, and linking is strong only when the candidate space is restricted. The consistent message is that evaluation design, not model capability alone, drives the reported numbers.

### 4.1 Strengths and Limitations

This study should be read in light of several limitations.

#### Translated, not native, evaluation data

The Dutch and Italian results are obtained on machine-translated text with projected annotations rather than native clinical notes, so they cannot, on their own, establish performance on native non-English text. This is a deliberate design choice: in the absence of natively annotated disorder corpora for these languages, annotation-preserving translation is an established and validated strategy [13] that enables a controlled, like-for-like cross-lingual comparison, and the near-complete annotation projection (about 0.98–0.99) indicates that the labels transfer faithfully. The method was validated for Dutch; its extension to Italian and a human expert review are natural next steps rather than threats to the present conclusions.

#### Potential circularity

Because the same model family produced the translations and the extractions, and the corpus derives from the public MIMIC database, agreement could in principle be inflated by memorisation. We tested this directly rather than leaving it open: an open-weight extractor (GLiNER) run on the same text scored far lower (overlap F1 0.11–0.19), and performance remains well below the supervised SemEval-2014 Task 7 system, both indicating that the results reflect genuine extraction rather than a generic artefact of the translated text. A residual effect cannot be entirely excluded, as in any work on publicly derived corpora, but the evidence suggests it is limited.

#### Sensitivity to evaluation design

Headline scores depend on the matching criterion and the candidate space, as they do throughout the clinical NLP literature. Rather than report only the most favourable setting, we provide both permissive and strict matching and both the restricted and the realistic candidate space, so that the deployment-relevant numbers are explicit; we regard this transparency as a feature of the evaluation rather than a shortcoming.

#### Scope

The evaluation covers one corpus, one LLM (GPT-4o), and the disorder semantic group. This focus is shared by much of the field and matches the canonical ShARe/CLEF and SemEval benchmarks, in which disorders are the central entity type, and GPT-4o is a representative strong general-purpose LLM. It is, however, proprietary and closed-weight, which constrains exact reproducibility. We mitigate this by releasing our predictions and evaluation code and by reporting an open-weight (GLiNER) reference point. We delimit our claims accordingly, and the released code makes extension to further entity types, corpora, and models straightforward.

#### Clinical use

As for any clinical NLP system, post-processing and human oversight remain necessary before deployment [35, 37]; this is a general prerequisite for the field rather than a shortcoming specific to our pipeline.

This is one of the first systematic evaluations of generative LLMs for multilingual clinical NER and BEL across English, Dutch, and Italian. The annotation-preserving translation pipeline provides a reproducible and scalable strategy to extend English corpora to low-resource languages, facilitating cross-lingual research [13]. By combining generative LLMs for entity recognition with embedding-based models for entity linking, we show a modular architecture adaptable to diverse data availability and resource constraints. The zero-shot setup reduces reliance on costly annotated corpora, enabling rapid deployment in new languages or domains [16, 41]. Our evaluation framework is comprehensive, covering entity recognition, annotation projection, and linking performance under both permissive and strict criteria. Finally, the error analysis showed that GPT sometimes identified clinically relevant concepts, suggesting that LLM-based extraction can complement human curation and improve dataset completeness [35, 38].

### 4.2 Future Work

Future research should extend our analysis beyond GPT-4o to include open-weight architectures and domain-adapted models, in order to better understand how model design and training resources influence multilingual clinical NLP performance [20,42]. Another direction is the systematic exploration of strategies to improve performance, such as refined prompt engineering, few-shot in-context learning, and integration of domain knowledge through constrained decoding or external knowledge sources [41]. Post-processing remains equally promising: re-ranking entity linking candidates with LLMs or ontology-aware methods may reduce ranking ambiguities and improve normalization accuracy [21, 39].

Another avenue lies in generating standardized outputs that can be added to data models such as OMOP CDM, thereby facilitating interoperability and downstream research applications [43]. We also plan to investigate language-specific documentation styles and terminological variability to adapt prompts and post-processing steps to local practices, reducing performance gaps across languages. Expanding the multilingual coverage of biomedical ontologies such as UMLS is critical to ensure equitable clinical NLP beyond English [39].

Finally, evaluating the pipeline on real-world clinical data will allow assessment of robustness and generalizability beyond benchmark corpora. By addressing these points, future systems may achieve greater reliability and clinical utility across diverse languages and care settings.

## 5 Conclusions

Across English, Dutch, and Italian, a zero-shot LLM pipeline for clinical disorder recognition and linking partially replicates a human annotator without any task-specific training. Recognition is moderate and well below supervised systems, and its apparent quality depends strongly on evaluation design (overlap versus strict matching, restricted versus realistic linking). The pipeline is conservative and high-precision; in English it is competitive with established clinical taggers, and its distinguishing value is that the same pipeline transfers unchanged to Dutch and Italian, where such tools are scarce. Many of its apparent errors are real clinical concepts rather than hallucinations, suggesting a role in supporting, not replacing, human curation. Because the Dutch and Italian results are evidence on translated benchmark text, future work should validate on native non-English clinical notes, extend beyond disorders, and compare against trained clinical systems before broader claims about multilingual clinical information extraction are made.

## Data Availability

The data used in this study are available upon reasonable request from the ShARe/CLEF data providers and are subject to their data access policies.

## Declarations

### Ethics approval and consent to participate

The corpus is derived from de-identified clinical reports from the MIMIC database, which has institutional review board (IRB) approval for research use. All patient data used in this study were fully anonymized prior to use, and access was governed by the relevant data use agreement. No additional ethical approval was required for this secondary analysis of existing de-identified data.

### Consent for publication

Not applicable.

### Availability of data and materials

The underlying ShARe corpus (SemEval-2014 Task 7) derives from the MIMIC database and is governed by a PhysioNet data use agreement, so the translated Dutch and Italian corpora cannot be redistributed. To support reproducibility, all code for the translation, annotation projection, entity extraction, linking, and evaluation is publicly available at https://github.com/saramazz/llm_EMC; users who hold the relevant licences can regenerate the translated corpora from the original ShARe data with these scripts.

### Authors’ contributions

S. Mazzucato: conceptualization, methodology, software, validation, formal analysis, investigation, data curation, writing (original draft), writing (review and editing), visualization. T.M. Seinen: conceptualization, methodology, resources, writing (review and editing), supervision. S. Moccia: writing (review and editing), supervision. S. Micera: writing (review and editing), supervision, funding acquisition. A. Bandini: writing (review and editing), supervision. E.M. van Mulligen: resources, writing (review and editing), supervision. All authors read and approved the final manuscript.

## Acknowledgements

The authors thank the “Proximity Care Project,” led by Scuola Superiore Sant’Anna, Interdisciplinary Center “Health Science” with support from Fondazione Cassa di Risparmio di Lucca, and conducted in collaboration with local healthcare and municipal partners in the Lucca province. The authors also acknowledge the Department of Medical Informatics at Erasmus University Medical Center for their support.

---

**Additional file 1.** Supplementary material (PDF): the full clinical entity-recognition prompt (Section S1), NER error examples (S2), recall by UMLS semantic type with support counts (S3), full biomedical entity linking results (accuracy@1, accuracy@3, accuracy@5 and MRR, restricted and realistic; S4), and the QuickUMLS threshold sweep (S5).

