## Supplementary material for "Advancements in Multilingual Biomedical Natural Language Processing: Exploring Large Language Models for Named Entity Recognition and Linking"

Code and saved predictions: https://github.com/saramazz/llm_EMC

---

### S1. Full Prompt for Clinical Entity Recognition

The text below is the exact prompt used to produce the GPT annotations, reproduced verbatim and applied unchanged to English, Dutch, and Italian (temperature 0.5, max_tokens 2000). At inference the clinical note to be annotated was appended after this prompt, enclosed between triple quotes (''' ... ''').

```
The goal of this task is to review clinical notes, identify mentions of disorders and
relevant additional information, and annotate them directly in the provided text without
altering any other content.

Instructions:
1. Preserve the Complete Original Text: Maintain the exact wording, typos, and formatting
, including the total character count. Do not omit any part of the text provided between
''' '''. In the answer do not report the ''' '''.
2. Annotation Integration: Insert annotations within double brackets [[]] directly around
 relevant disorder mentions in the original text. Ensure the original structure and
sequence of text remain unchanged.
3. Full Text Output: Return the entire input text with inserted annotations. Do not
provide separate lists or summaries.
4. No Corrections: Retain all typographical errors, unusual formatting, and punctuation
as they appear in the source text.
5. Bracket Annotations Only: The sole modification allowed is the addition of brackets
for annotation|no other text modifications are permitted.

Disorder Categories (for annotation reference):
- Congenital Abnormality
- Acquired Abnormality
- Injury or Poisoning
```

- Pathologic Function
- Disease or Syndrome
- Mental or Behavioral Dysfunction
- Cell or Molecular Dysfunction
- Experimental Model of Disease
- Anatomical Abnormality
- Neoplastic Process
- Signs and Symptoms

Annotation Guidelines:

1.Identifying Disorder Mentions
Annotate disorder mentions that explicitly describe a condition, symptom, or diagnosis.
Example
Input: The patient reported left-sided facial droop.
Output: The patient reported left-sided [[facial droop]].
Input: The patient has a history of hypertension.
Output: The patient has a history of [[hypertension]].

2. Use of Synonyms
Annotate terms that are reasonable synonyms of a disorder, even if the exact phrase differs or if they are disjoint.
Example:
Input: The patient reported left-sided facial droop.
Output: The patient reported left-sided [[facial droop]].
Input: His face was weak.
Output: His [[face ... weak]].

3. Explicit Mentions Only
Only annotate disorders explicitly stated in the text.
Do not annotate inferred conditions.
Example:
Input: The patient's chem profile reveals a 138 sodium, 4.0 potassium, 106 chloride, 19 bicarb, 43 and 2.2 are the BUN and creatinine, indicating probable blood in the GI tract.
Output: The patient's chem profile reveals a 138 sodium, 4.0 potassium, 106 chloride, 19 bicarb, 43 and 2.2 are the BUN and creatinine, indicating probable [[blood in the GI tract]].
Input: She had a Crohn flare with symptoms of bowel obstruction that typically resolves with rehydration. Her current symptoms are reminiscent of this.
Output: She had a [[Crohn]] flare with symptoms of [[bowel obstruction]] that typically resolves with rehydration.
Example (No Annotation Needed):
Input: The lab results indicate an elevated white blood cell count, suggesting an infection.

Output: The lab results indicate an elevated white blood cell count, suggesting an infection.

4. Maintain Specificity
Only annotate the most specific disorder mentioned in the text.
Example:
Input: The patient has a small bowel obstruction.
Output: The patient has a [[small bowel obstruction]].
Input: The patient was found to have left lower extremity DVT.
Output: The patient was found to have left [[lower extremity DVT]].
Input: The patient has severe pre-eclampsia.
Output: The patient has [[severe pre-eclampsia]].
Input: The patient has chronic gingivitis.
Output: The patient has [[chronic gingivitis]].

5. Negation and Temporality
Do not include negations or temporal aspects within disorder annotations.
Example:
Input: No chest pain was reported.
Output: No [[chest pain]] was reported.
Input: No evidence of myocardial infarction.
Output: No evidence of [[myocardial infarction]].
Input: The patient has a history of stroke.
Output: The patient has a history of [[stroke]].
Input: The patient had a heart attack five years ago.
Output: The patient had a [[heart attack]] five years ago.

6. Independence from Syntactic Structure
A disorder mention can span noun phrases or other syntactic units.
Example:
Input: The patient had a tumor of the skin removed.
Output: The patient had a [[tumor of the skin]] removed.
Input: A tumor was found in the left ovary.
Output: A [[tumor]] was found in the left [[ovary]].

7. Not Patient-Specific
Annotate all disorder mentions, regardless of whether they refer to the patient or another person.
Example:
Input: The patient's father has schizophrenia.
Output: The patient's father has [[schizophrenia]].
Input: He should return to the ED immediately if any rash occurs.
Output: He should return to the ED immediately if any [[rash]] occurs.

```
Input: The patient was referred to the lupus clinic.
Output: The patient was referred to the [[lupus]] clinic.

8. All Disorder Mentions Must Be Annotated
Every disorder mention must be annotated, no matter where it appears in the note.
Example:
Input: He has diabetes, hypertension, and chronic kidney disease.
Output: He has [[diabetes]], [[hypertension]], and [[chronic kidney disease]].



Here is the clinical note text to be annotated:
```

---

### S2. NER Error Examples

##### Category 1: Missed Extractions

- **Heroine use:** Clinicians annotated “[[heroine use]]” in Past Medical History, but GPT marked it as “[[heroin use]]” and in some parts omitted it.
- **Overdose (considered):** Clinicians annotated possible “[[overdose]]” in HPI and Hospital Course. GPT only captured it once, missing the second occurrence.
- **Toxicity:** Clinicians annotated “[[toxicity]]” as part of differential. GPT dropped this annotation.
- **Clamped jaw / twitching:** Clinicians annotated “[[clamped jaw]]” and “[[twitching]]” during seizure-like activity. GPT captured seizure but omitted these finer details.
- **Decerebrate posturing:** Clinicians annotated “[[decerebrate posturing]]”. GPT omitted it.

##### Category 2: Entities Extracted Only by GPT

- **Zofran:** GPT annotated “[[Zofran]]”, but clinicians did not mark it (original text: “There she was given Zofran and Ativan”).
- **Ativan:** GPT annotated “[[Ativan]]” as an entity, while clinicians only mentioned it without annotation (same sentence as above).

- **Mucomyst:** GPT annotated "[[Mucomyst]]"; clinicians left it unannotated.
- **Hysterectomy:** Original text: "she had a hysterectomy in July". GPT annotated "[[hysterectomy]]", although clinicians did not.
- **C-section / D&C:** GPT annotated "[[c-section]]" and "[[D&C]]"; clinicians listed them but did not annotate.
- **MVI / folate / thiamine:** GPT annotated these supplements; clinicians did not.
- **Weight loss:** Original text stated "not having experienced any weight loss". GPT incorrectly annotated "[[weight loss]]" as a positive finding, hallucinating a condition.
- **Distress:** Original text: "in no acute distress". GPT hallucinated "[[distress]]" as present.
- **Lesions:** Original text: "no lesions". GPT annotated "[[lesions]]" as a positive finding.
- **Lymphadenopathy:** Original text: "no lymphadenopathy". GPT annotated "[[lymphadenopathy]]" as positive.
- **Masses:** Original text: "no palpable masses". GPT annotated "[[masses]]".
- **Abdomen:** Original text: "His abdomen was soft...". GPT annotated "[[abdomen]]" as a clinical concept, though it was only descriptive.
- **Guarding / rebound:** Both negated in the text ("No guarding", "No rebound"), but GPT annotated them as positive.
- **Pulmonary deterioration:** Original text described "some degree of pulmonary deterioration"; GPT hallucinated a non-standard medical entity with a placeholder code.
- **Ivor-Lewis esophagectomy:** GPT marked "[[Ivor-Lewis esophagectomy]]", while the gold annotation contained a de-identified broken string ("Ivor-[Doctor Last Name] esophagectomy"), illustrating inconsistent handling of de-identified text.

These examples illustrate the quantitative error pattern reported in the main text: in English most false positives are isolated non-disorder concepts (procedures, medications) or negated findings tagged as present, while genuine hallucinations (no matching UMLS concept) are a minority.

---

### S3. NER Recall by UMLS Disorder Semantic Type

Table S1 reports GPT-4o recall by UMLS semantic type on the CUI-bearing gold under overlap matching, with the number of gold mentions ($n$) per group. Recall is consistently lower in English than in Dutch and Italian across all types; signs/symptoms, pathologic functions and findings are recovered least reliably, whereas diseases/syndromes and neoplasms are recovered best. Precision and F1 are not defined per type because false positives cannot be assigned a gold semantic type.

| | | **English** | | **Dutch** | | **Italian** | |
|---|---|---|---|---|---|---|---|
| **Semantic type** | **TUI** | $n$ | R | $n$ | R | $n$ | R |
| Disease or Syndrome | T047 | 2541 | 0.73 | 2514 | 0.77 | 2534 | 0.78 |
| Sign or Symptom | T184 | 1396 | 0.43 | 1361 | 0.59 | 1395 | 0.60 |
| Pathologic Function | T046 | 896 | 0.37 | 877 | 0.51 | 888 | 0.57 |
| Neoplastic Process | T191 | 143 | 0.72 | 138 | 0.83 | 143 | 0.76 |
| Mental/Behavioral Dysfunction | T048 | 170 | 0.65 | 167 | 0.78 | 171 | 0.74 |
| Injury or Poisoning | T037 | 265 | 0.39 | 258 | 0.57 | 263 | 0.53 |
| Finding | T033 | 247 | 0.28 | 241 | 0.43 | 239 | 0.51 |
| Other | — | 194 | 0.44 | 193 | 0.51 | 193 | 0.54 |
| All CUI-bearing | — | 5956 | 0.56 | 7758 | 0.56 | 5938 | 0.67 |

Table S1: GPT-4o NER recall by UMLS disorder semantic type (overlap matching, CUI-bearing gold; 131-document evaluation set). The "All CUI-bearing" row for Dutch equals the full-gold recall (0.56) because Dutch has no CUI-less mentions; per-type rows cover the TUI-mapped subset (5,857 of 7,758 Dutch gold mentions).

---

### S4. Full Biomedical Entity Linking Results

Table S2 reports accuracy@1, accuracy@3, accuracy@5 and mean reciprocal rank (MRR) for all three encoders in both the restricted (corpus-CUI) and realistic (semantic-type-filtered UMLS) candidate spaces, per language. All encoders are near ceiling in the restricted setting; performance drops sharply in the realistic setting, where multilingual SapBERT is the most reliable. The realistic accuracy is bounded above by the candidate-coverage ceiling (some gold CUIs fall outside the candidate set). On the linkable subset (excluding CUI-less gold), multilingual SapBERT reaches accuracy@1 of 0.77 (English), 0.63 (Dutch), and 0.64 (Italian).

---

| Encoder | Setting | acc@1 | acc@3 | acc@5 | MRR |
|---|---|---|---|---|---|
| *English* | | | | | |
| all-MiniLM-L6-v2 | restricted | 0.94 | 0.97 | 0.97 | 0.96 |
| | realistic | 0.69 | 0.76 | 0.76 | 0.72 |
| BioBERT | restricted | 0.94 | 0.96 | 0.97 | 0.95 |
| | realistic | 0.26 | 0.36 | 0.41 | 0.32 |
| mSapBERT | restricted | 0.94 | 0.97 | 0.97 | 0.95 |
| | realistic | 0.69 | 0.76 | 0.77 | 0.72 |
| *Dutch* | | | | | |
| all-MiniLM-L6-v2 | restricted | 0.91 | 0.94 | 0.95 | 0.93 |
| | realistic | 0.56 | 0.61 | 0.62 | 0.59 |
| BioBERT | restricted | 0.91 | 0.92 | 0.92 | 0.91 |
| | realistic | 0.49 | 0.55 | 0.56 | 0.52 |
| mSapBERT | restricted | 0.91 | 0.95 | 0.96 | 0.93 |
| | realistic | 0.63 | 0.67 | 0.68 | 0.65 |
| *Italian* | | | | | |
| all-MiniLM-L6-v2 | restricted | 0.90 | 0.96 | 0.96 | 0.93 |
| | realistic | 0.52 | 0.58 | 0.59 | 0.55 |
| BioBERT | restricted | 0.90 | 0.93 | 0.94 | 0.92 |
| | realistic | 0.35 | 0.41 | 0.44 | 0.38 |
| mSapBERT | restricted | 0.91 | 0.96 | 0.96 | 0.93 |
| | realistic | 0.56 | 0.61 | 0.62 | 0.59 |

Table S2: Biomedical entity linking: accuracy@1, accuracy@3, accuracy@5 and MRR by encoder, candidate setting, and language (full mention set). In the realistic setting accuracy@1 is bounded above by the candidate-coverage ceiling (0.85 English, 0.88 Dutch, 0.77 Italian), since some gold CUIs fall outside the candidate set. mSapBERT = multilingual SapBERT (`SapBERT-UMLS-2020AB-all-lang-from-XLMR`).

### S5. English NER Baselines: QuickUMLS Threshold Sensitivity

Table S3 reports QuickUMLS performance on the English test set as a function of its Jaccard similarity threshold (disorder semantic types only, scored against the same gold as GPT-4o). The default threshold (0.70) over-generates (low precision); F1 rises with a stricter threshold. In the main text we report QuickUMLS at the default 0.70 and at 0.80; for completeness, the best operating point (0.95) reaches overlap F1 0.68. MedCAT (`umls_sm`, MIMIC-III trained) reached overlap F1 0.63 (precision 0.55, recall 0.74), strict F1 0.54, and concept accuracy@1 0.75; GPT-4o reached overlap F1 0.60 and strict F1 0.49 (see main Table 3).

| Threshold | P | R | F1 (overlap) | F1 (strict) | CUI@1 |
|---|---|---|---|---|---|
| 0.70 (default) | 0.43 | 0.68 | 0.52 | 0.34 | 0.66 |
| 0.80 | 0.63 | 0.65 | 0.64 | 0.51 | 0.74 |
| 0.95 | 0.75 | 0.62 | 0.68 | — | — |

Table S3: QuickUMLS English NER performance versus similarity threshold (overlap and strict span matching), disorder semantic types.
